# Language-dependent diagnostic safety of medical AI systems: a cross-lingual benchmarking and prospective clinical study

**DOI:** 10.64898/2026.05.19.26353490

**Authors:** Yuqian Wang, Hongyu He, Rongpeng Zhu, Yunyi Lu, Pawit Phadungsaksawasdi, Manqiang Peng, Zengping Liu, Ke Zou, Ye Zhang, Sien Ping Chew, Yih Chung Tham, Arian Khorasani, Hao Deng, Ching-Yu Cheng, Jie Yang, Dianbo Liu

## Abstract

**Background:** Patients worldwide receive healthcare in many languages, yet medical AI systems are validated almost exclusively in high-resource languages such as English and Chinese, exposing patients in other linguistic settings to unquantified diagnostic risk. Existing multilingual evaluations rely on translated research-style benchmarks that fail to capture authentic clinical work. We aimed to characterise the patient safety consequences of multilingual medical AI deployment in real-world clinical settings and to develop an auditable detection method for unsafe outputs.

**Methods:** We evaluated different language models(LLMs) and visual language models(VLMs) across four real-world clinical tasks (conversational QA, radiology report generation, glaucoma diagnosis, ICU re-intubation prediction) in five languages (English, Chinese, Malay, Thai, Persian). We developed a token-level uncertainty toolkit to localise reasoning instability, com pared three inference paradigms (native-language, English chain-of-thought, back-translation pivot), and conducted a prospective study (50 dialogues, 150 physician-reviewed records).

**Findings:** LLMs/VLMs performance degraded consistently from high-to low-resource languages across al l tasks. Key gaps included: HealthBench score declining from 0·3743 to 0·3180; radiology macro-F1 from 0·2938 to 0·2149–0·2424, consistent with selective pathology suppression; glaucoma accuracy from 50·7% to 32·7%; ICU parameter recall from 100·0% to 48·5%. Multimodal inputs amplified degradation. Qwen3 VL 235B showed attenuated decline with no re source-ordered pattern in glaucoma classification. Token-level analysis localised instability to mid-chain stages (40–70% of the normalised trajectory); perplexity-based confidence failed to flag errors (AUROC 0·41–0·66). Back-translation pivot consistently restored performance. In the prospective study, 98·7% of records required physician edits (overall modification score 53·6%); Thai-pivot correction burden (59·0%) exceeded English-pivot (5 0·7%, p=0·003) and Chinese-direct (51·0%, p=0·004).

**Interpretation:** Multilingual deployment produced clinically consequential failures, including missed pathology, distorted physiological extraction, and amplified multimodal misclassification, that were invisible to monolingual validation and not reliably flagged by model confidence. Pre training data composition may contribute to multilingual safety risk. Language-specific safety auditing should precede deployment in non-dominant-language healthcare settings; the open-source detection toolkit enables this without model retraining.

## Introduction

A substantial proportion of the global population receives healthcare in linguistically diverse settings, including in languages that are underrepresented in current biomedical AI development and evaluation. Health systems across South-East Asia, the Middle East, and sub-Saharan Africa are increasingly adopting AI-assisted tools for clinical decision support, radiology reporting, and diagnostic classification.^1–3^ Yet system validation remains overwhelmingly using high-resource languages such as English and Chinese,^4^ and we know little about the real-world safety of these systems for the patients most likely to encounter the m outside research settings. Cross-lingual performance gaps are common across general AI applications.^5,6^ In clinical AI, they directly affect diagnostic interpretation, risk stratification, and treatment decisions.^7^ Such failures are often subtle: models produce clinically plausible but systematically incomplete or incorrect outputs (missed pathology in a chest radiograph report, misclassified glaucoma severity, misreported ICU physiological parame ters) that escape detection during routine use.^18^

Prior multilingual evaluations of medical AI have reported cross-lingual performance differences, with healthcare-query studies suggesting English prompting can outperform non-English prompting.^8^ However, these studies remain limited in three aspects. First, most studies use benchmarks built from multiple-choice questions rather than real-world clinical tasks. Aggregate accuracy metrics hide failure modes that matter clinically, such as selective suppression of pathological findings. Second, evaluations have largely been text-only, leaving multimodal settings (radiology, ophthalmology, ICU monitoring) unexamined. In these set tings, visual and linguistic reasoning must be jointly processed, and failures may be amplified. Third, no study has systematically tested whether model confidence reliably flags diagnostic errors across languages,^9^ a precondition for any safety filter to protect patients in multilingual deployment. Recent work has shown that requiring large reasoning models to think in non-English languages degrades accuracy,^10^ indicating a trade-off between native-language reasoning and performance. As medical large language models (LLMs) and vision-language models (VLMs) move from prototypes to clinical deployment, these gaps represent an unquantified patient safety risk.^17^

To characterise these risks, we evaluated multiple LLMs and VLMs across four clinical task domains and five languages spanning high-to low-resource representation. To detect clinic ally risky outputs that conventional confidence scores fail to flag, we developed a token-level uncertainty analysis method that localises reasoning instability during inference, building on prior work identifying reasoning as both a central capability and a vulnerability of LLMs.^11,12^ We released this method as an open-source safety auditing toolkit (web inter face: https://doctor-language-bridge.onrender.com/; source code: https://github.com/wang-ava/languagegap), which lets clinicians submit clinical questions in any language and visualise where diagnostic uncertainty concentrates along the translation–reasoning–back-translation pipeline. We further compared three inference paradigms (native-language reasoning, English chain-of-thought, and back-translation pivot) and identified back-translation a s a training-free mitigation that often improved performance. To extend these findings to clinical practice, we conducted a prospective study in an ophthalmology outpatient clinic, testing whether benchmark failure patterns persist in real-world use. Together, our findings provide empirical evidence for pre-deployment safety assessment of clinical AI systems serving non-English-speaking patient populations, who currently bear an under-monitored share of multilingual diagnostic risk.^19^

## RESEARCH IN CONTEXT

### Evidence before this study

We searched PubMed and Google Scholar through May 2026 for studies on multilingual clinical AI performance, without language restrictions. Existing evaluations have predominantly used translated QA benchmarks with aggregate scoring. Cross-lingual performance gaps are established in general NLP, but whether they lead to missed diagnoses, selective loss of safety-critical findings, or false reassurance through uncalibrated confidence has not been systematically studied. Multimodal clinical tasks such as radiology and ophthalmic diagnosis have received particularly limited multilingual scrutiny. Language-specific safety testing is not yet standard in clinical AI deployment.

### Added value of this study

We evaluated multilingual safety of medical LLMs and VLMs across four clinical domains: medical QA, EHR-based glaucoma diagnosis, radiology report generation, and ICU re-intubation prediction. Degradation was systematic and resource-ordered. In radiology, deficits concentrated on pathological finding detection while overall accuracy was preserved, revealing a false-negative bias invisible to aggregate metrics. Token-level analysis localised uncertainty to mid-chain reasoning stages. Perplexity-based confidence failed to separate correct from incorrect outputs. A training-free back-translation pivot consistently improved performance, suggesting reasoning instability rather than absent medical knowledge, though the mechanism is unproven. We validated these findings prospectively: a blinded attending ophthalmologist reviewed 150 AI-generated records from three pipelines applied to 50 de-identified dialogues (from 165 source recordings). Degradation persisted in authentic clinical speech. We introduced a modification-score framework decomposing physician correction burden by pipeline, field, edited-field count, and edit type; most edits were substantive clinical corrections. Paired dialogue-level comparisons separated pipeline-specific effects from case mix.

### Implications of all the available evidence

English-centric validation creates a blind spot allowing dangerous AI failures to reach patients undetected. Multilingual degradation can manifest silently, through reduced pathology detection and degraded multimodal classification, without drops in model confidence. Pre-deployment evaluation should include language-specific safety auditing, particularly for systems serving linguistically under-represented populations. Regulatory frameworks should consider requiring multilingual performance equivalence evidence. Back-translation may reduce risk but requires prospective validation. In our prospective study, the near-universal edit rate of 98·7% confirms that human oversight remains necessary.

## Methods

### Study design

In this study, we conducted multiple real-world cross-lingual clinical benchmarking tasks and a prospective study. The benchmark component tested language-dependent diagnostic performance under controlled conditions. The prospective study tested whether the same risk appeared in real outpatient documentation. English was the source and reference condition. Chinese, Malay, Thai, and Persian were selected to span high-to low-resource language representation. The four tasks were chosen to cover common clinical AI use cases: free-text medical advice, radiology report generation, ophthalmology diagnosis, and ICU risk prediction. This design allowed us to test whether language effects repeated across clinical formats.

The benchmark datasets were HealthBench conversational medical question answering,^13^ an institutional OphthalmologyEHR-Glaucoma dataset, MIMIC-CXR v2·0·0,^14^ and MIMIC-III.^15^ HealthBench tested open-ended clinical advice. MIMIC-CXR tested image-to-report generation. OphthalmologyEHR-Glaucoma used two input configurations: 2type, which includes textual case descriptions and tabular data, and 3type, which additionally incorporates fundus photographs. MIMIC-III tested extraction and interpretation of physiological data for re-intubation risk. GPT-5.2 was used for HealthBench after GPT-4o produced near-zero non-English scores under the strict rubric, prompting a shift to GPT-5.2 due to its stronger multilingual and medical reasoning capabilities. GPT-4o was still used for the other three datasets, while in HealthBench it was retained solely for token-level analysis, as it provided detailed token -level outputs. Qwen3 VL 235B A22B Thinking was included as a multilingual reference model. We tested three reasoning strategies: native-language reasoning, where both reasoning a nd response were conducted entirely in the target language; English chain-of-thought, where permitted, which forced the reasoning process to be in English while keeping the final answer in the target language; and a back-translation pivot, where the target-language input was first translated into English before reasoning and answering. Reporting followed TRI POD+AI where applicable.^16^ Detailed model-selection rationale and inference-paradigm applicability are provided in Appendix 1.

**Table 1.**
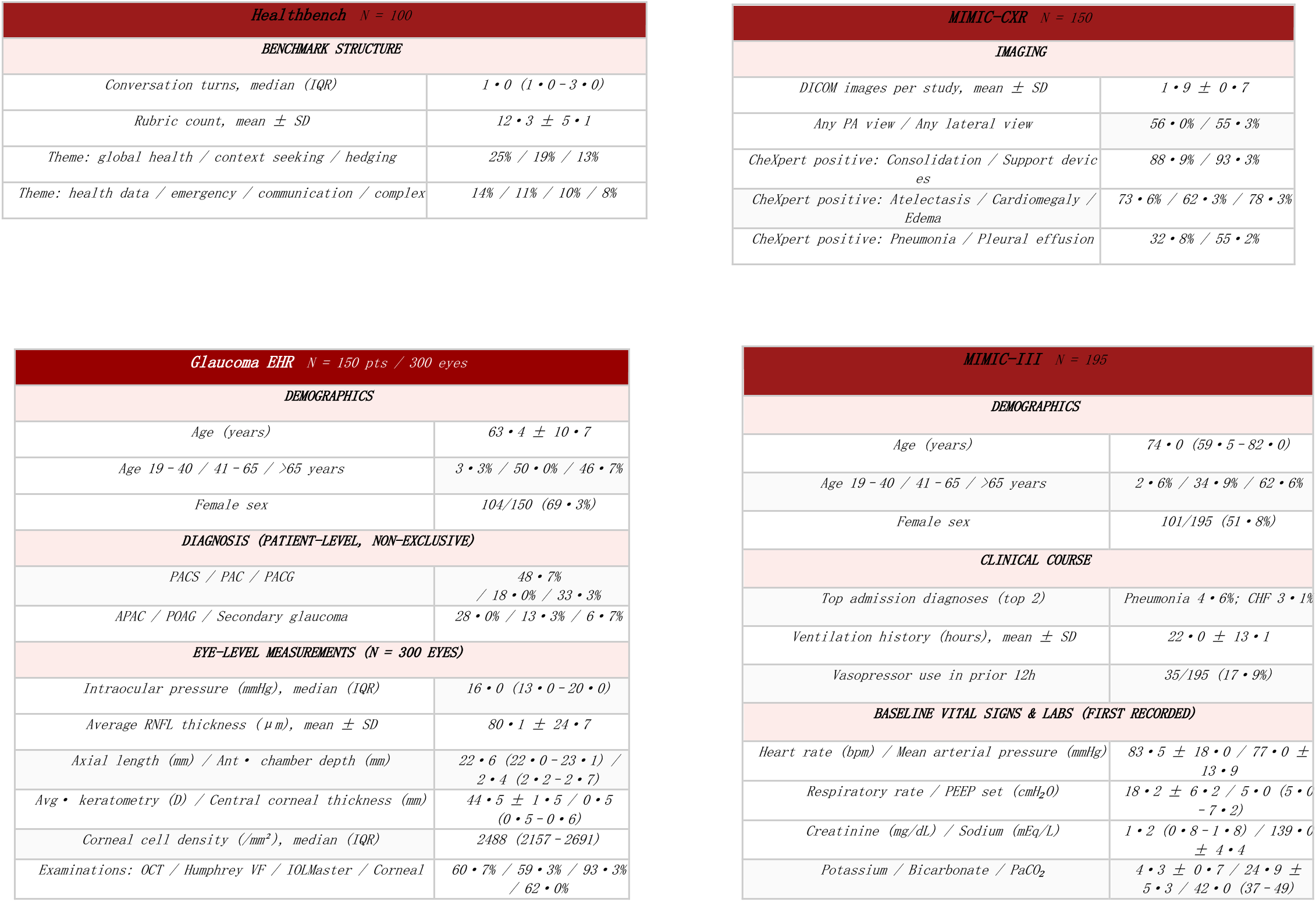
Descriptive Characteristics of the Four Study Datasets. Continuous variables: mean ± SD or median (IQR). Categorical variables: n/N (%) or % where space-condensed. PACS, Primary Angle Closure Suspect; PAC, Primary Angle Closure; PACG, Primary Angle Closure Glaucoma; APAC, Acute Primary Angle Closure; POAG, Primary Open-Angle Glaucoma; RNFL, Retinal Nerve Fibre Layer; VF, visual field; D, diopter; CHF, congestive heart failure; PEEP, positive end-expiratory pressure; PaCO_2_, partial pressure of carbon dioxide.

### Translation pipeline

We used a two-stage translation pipeline. English clinical inputs were translated into Chinese, Malay, Thai, and Persian with prompts that preserved medical terms, numbers, units, abbreviations, and negation. The prompts did not allow interpretation, summarisation, or explanatory additions. Clinicians reviewed forward translations for clinical accuracy, safe ty, and usability. Chinese and Malay translations reached near-ceiling ratings. Thai was lower but remained clinically usable. Persian remained high but below the ceiling. Back-translation metrics showed high preservation of clinically important content. Number F1 and N umber+Unit F1 exceeded 0·95, and Negation F1 ranged from 0·908 to 0·942.

Back-translation NLP metrics confirmed that clinically critical content was preserved with near-perfect fidelity: Number F1 and Number+Unit F1 exceeded 0·95 across all languages, and Negation F1 ranged from 0·908 to 0·942, confirming that numerical values and logical polarity, the two most consequential elements for clinical correctness, were not grossly distorted by the pipeline (Figure 2, right). Surface-level metrics showed expected lexical variation from legitimate paraphrasing rather than clear semantic loss. These checks reduce the chance that the main findings were caused by gross translation error, although subtler effects such as terminology standardisation or improved prompt-model alignment cannot be excluded.

**Figure 1.**
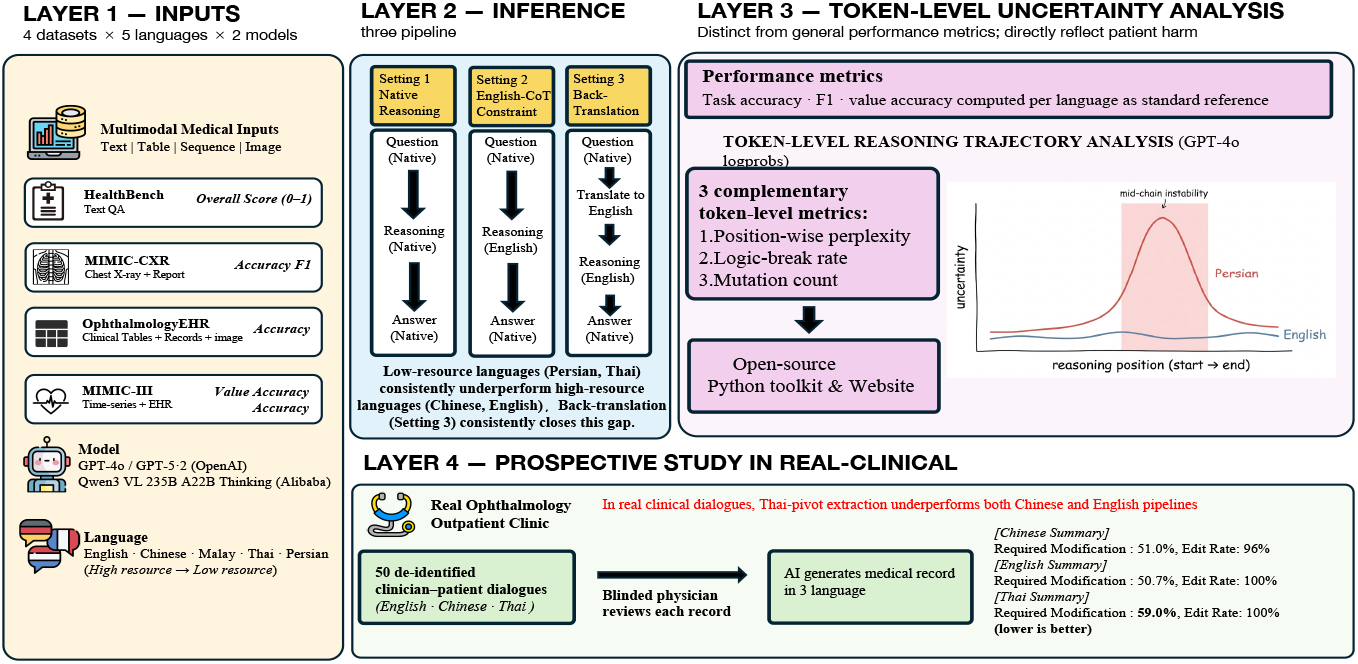
Overall study framework. Four medical datasets spanning text QA, radiology reporting, ophthalmological EHR, and ICU time-series are evaluated under three reasoning paradigms (Setting 1: native reasoning; Setting 2: English chain-of-thought constraint; Setting 3: back-translation pivot), yielding up to 13 outputs per sample (5 native-language responses across all languages, 4 English-CoT responses for non-English languages only, and 4 back-translation responses for non-English languages only). Benchmark findings are validated in a prospective study using 50 de-identified clinician–patient dialogues from a real-world ophthalmology outpatient clinic, with blinded physician review of 1 50 records across three pipeline variants (prospective study module, bottom panel).

**Figure 2.**
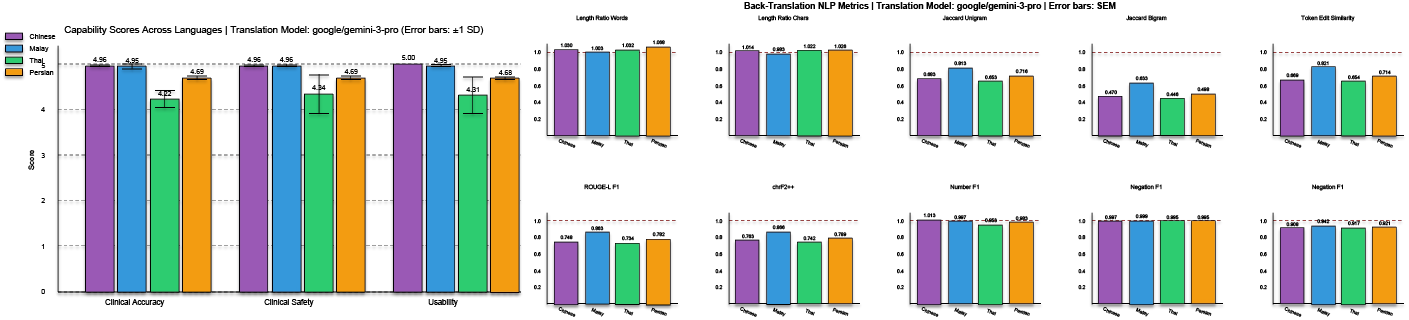
Forward-translation clinical review and back-translation fidelity of the two-stage translation pipeline (Google Gemini-3-Pro). Left: independent clinician ratings (1–5) of clinical accuracy, clinical safety, and usability for Chinese, Malay, Thai, and Persian forward translations. Right: back-translation NLP metrics including length ratios and surface-form similarity (Jaccard unigram/bigram, token edit similarity, ROUGE-L, chrF++), alongside clinically critical fidelity metrics (Number F1, Number+Unit F1, Negation F1). Dashed reference lines denote the original English value for metrics where applicable.

**Figure 3.**
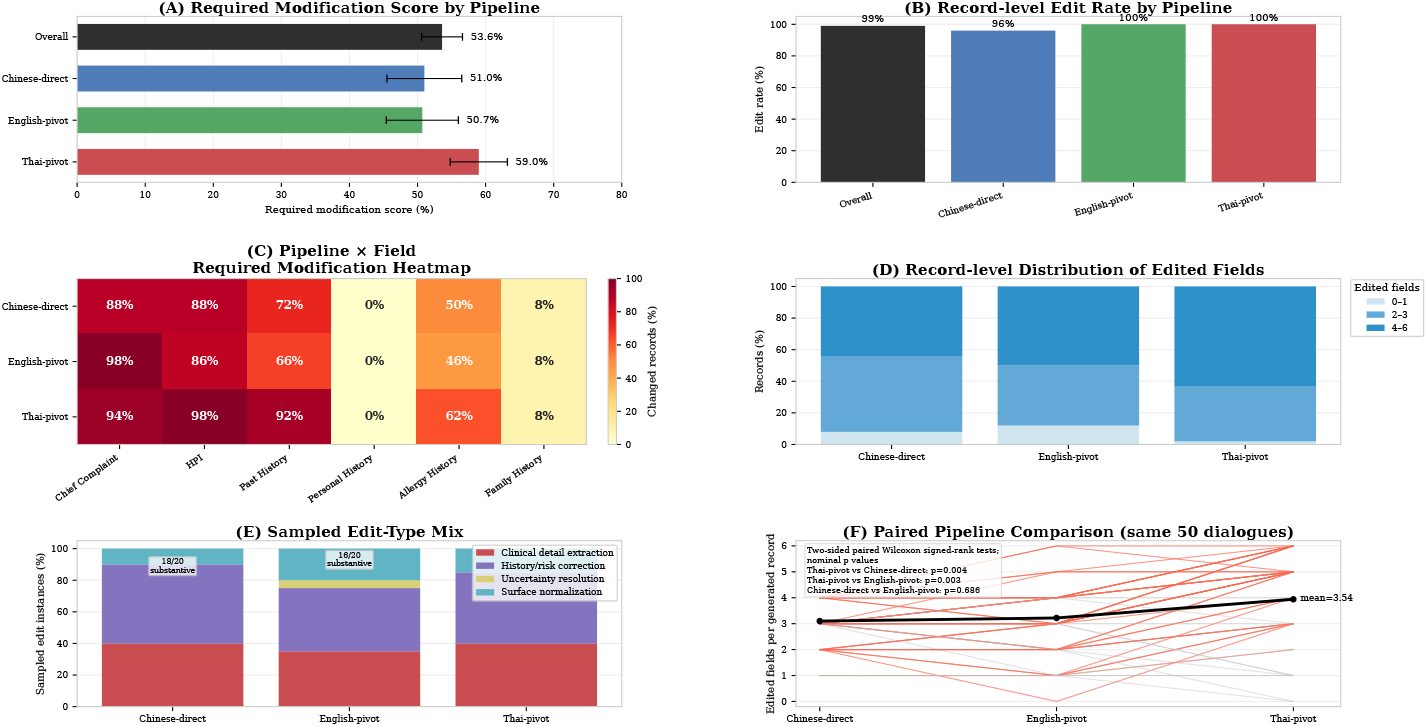
Physician correction burden across real-world multilingual documentation pipelines. Fifty de-identified ophthalmology outpatient dialogues were processed through Chinese-direct, English-pivot, and Thai-pivot pipelines, generating 150 structured records for blinded physician review. (A) Required modification score, defined as modified field slots divided by total editable field slots. Error bars indicate 95% CIs. (B) Record-level edit rate, defined as the proportion of records requiring at least one physician edit. (C) Field-level modification proportions across six documentation fields. Low modification in personal and family history should be interpreted with empty-field analysis because these fields often lacked extractable source information. (D) Distribution of edited-field counts per record. (E) Sampled edit-type mix, categorised as clinical detail extraction, history/risk correction, uncertainty resolution, or surface normalisation. Substantive edits were defined as the first three categories. (F) Paired dialogue-level comparison of edited-field counts across pipelines; pairwise comparisons used two-sided paired Wilcoxon signed-rank tests. HPI=history of present illness.

**Figure 4.**
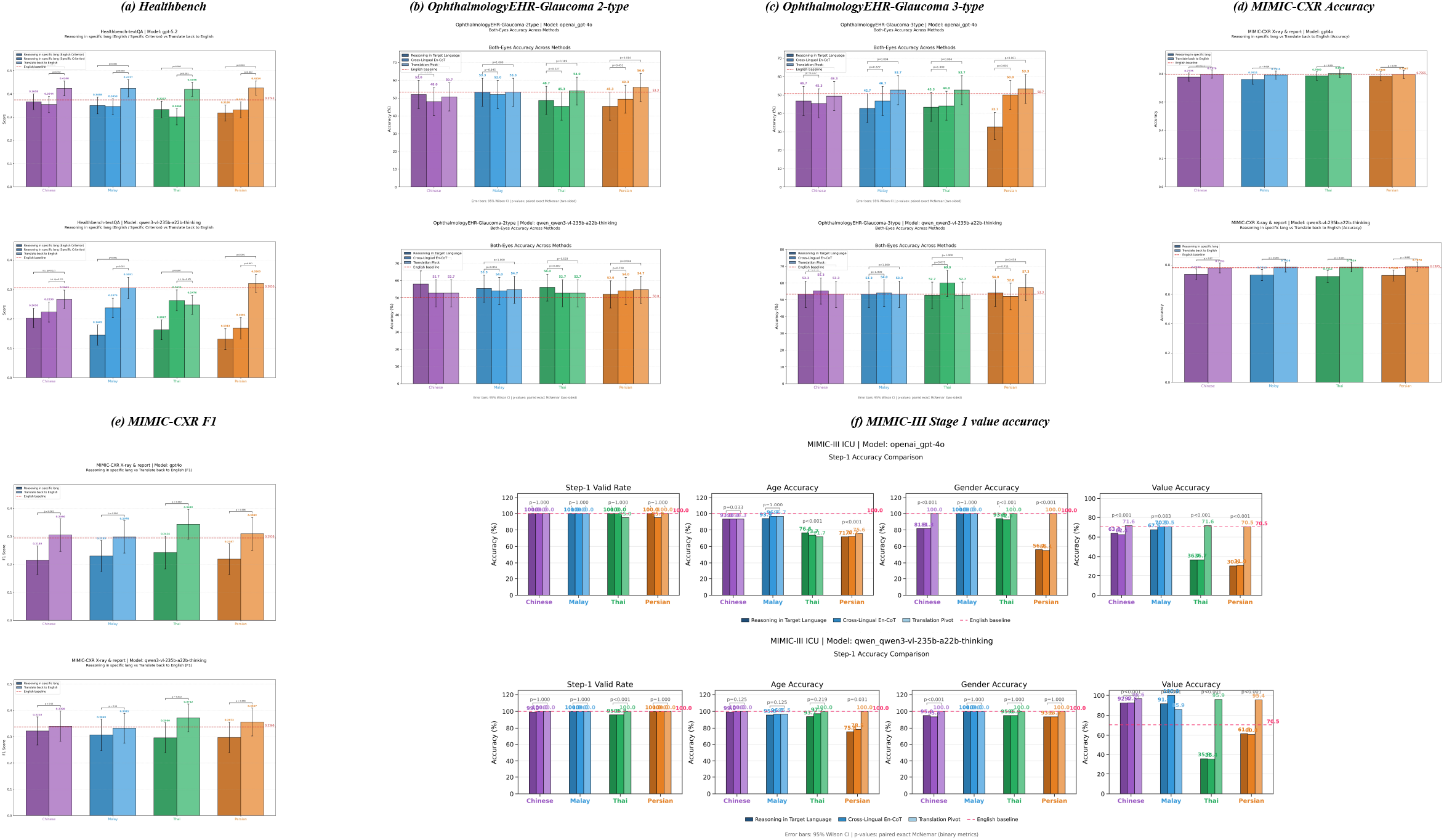
Cross-lingual performance across four medical benchmarks and three inference paradigms. (a) HealthBench text QA scores (GPT-5.2) for four non-English languages under native reasoning (Setting 1), English chain-of-thought constraint (Setting 2), and back-translation pivot (Setting 3); the red dashed line marks the English baseline. (b) OphthalmologyEHR-Glaucoma two-type accuracy. (c) Ophthalmology-EHR-Glaucoma three-type accuracy; the amplified gap relative to (b) illustrates super-additive multimodal degradation. (d) MIMIC-CXR accuracy across languages and settings. (e) MIMIC-CXR F1 score across languages and settings; note the accuracy–F1 dissociation under native reasoning. (f) MIMIC-III Stage 1 value accuracy, vital-sign parameter recall, and age accuracy under native reasoning (left) and back-translation (right). Back-translation (light bars) consistently recovered or exceeded the English baseline across all languages and benchmarks. CoT = chain of thought. F1 = harmonic mean of precision and recall

## Statistical and subgroup analysis

Primary endpoints were prespecified per dataset: rubric score (HealthBench), accuracy and macro-F1 (MIMIC-CXR), patient-level accuracy (OphthalmologyEHR-Glaucoma), actionable-parameter recall (MIMIC-III Stage 1). Cross-language differences were reported relative to English with bootstrap 95% CIs (10,000 resamples). No multiple-comparison correction was applied; pairwise differences were descriptive. Safety-relevant error patterns were also examined: missed pathological findings, actionable-parameter extraction failures, high-risk glaucoma misclassification.

Stratified analyses assessed whether multilingual degradation disproportionately affected key subgroups: acuity and complexity (HealthBench), urgent versus routine findings (MIMIC-CXR), high-risk and angle-closure subtypes (OphthalmologyEHR-Glaucoma), actionable versus overall parameters (MIMIC-III). Prospective scores were stratified by disease type, visit type, and record quality. (Definitions: Appendix 3; results: Appendix 10.)

### Behavioural analysis

Token-level log-probabilities were recorded. Three metrics were computed. Position-wise perplexity (exp(–log p)), normalised to 0–1 position and averaged across 20 bins, captured stage-dependent uncertainty. Logic-break rate was the proportion of samples with ≥1 token where adjacent log-probability drop > τ_drop=1·5 and either standardised drop > τ_z=2·0 or absolute value < τ_abs=–2·5. Mutation count tallied jumps ≥2·0 in absolute magnitude.

### Confidence calibration

Calibration was assessed for GPT-4o across OphthalmologyEHR-Glaucoma, MIMIC-III, language s, and inference settings. Confidence was windowed inverse perplexity over the middle 60% of tokens, avoiding template-token dominance. Metrics were AUROC, ECE, confidence separation gap, and top-bottom accuracy gap. Full definitions are provided in Appendix 2.

### Prospective study

The prospective clinical study was designed to test real clinical risk. We used 165 de-identified, audio-transcribed ophthalmology outpatient dialogues. Dialogues in Chinese dialects were quality-graded at source. High-quality dialogues had at least five turns with complete legible content. Low-quality dialogues had fewer turns or predominantly dialectal content. A stratified random sample of 50 dialogues preserved the natural mix of consultation quality, including dialect admixture. This design kept the prospective study close to clinic conditions and avoided selecting only clean transcripts.

Each dialogue was processed through three pipelines: Chinese-direct extraction, English-pivot extraction, and Thai-pivot extraction. Chinese-direct represents the source clinical language. English-pivot tested whether routing through English preserved or improved documentation quality. Thai-pivot tested whether routing through a lower-resource language increased correction burden. Non-Chinese outputs were back-translated into Chinese for blinded review. Each pipeline produced six editable fields: chief complaint, history of present illness, past medical history, allergy history, family history, and personal history.

An attending ophthalmologist reviewed all 150 generated records while blinded to pipeline assignment. The primary workflow endpoints were record-level edit rate and required modification score. The required modification score was the proportion of editable field slots that required physician correction. It captured the practical burden of making AI documentation usable. Paired dialogue-level comparisons used two-sided Wilcoxon signed-rank tests. Physician edits were also classified as surface normalisation, clinical detail extraction, history or risk correction, or uncertainty resolution. The last three categories were treated as substantive clinical edits. Sampling, field definitions, and modification-scoredet ails are provided in Appendix 8.

## Results

### Prospective study: multilingual degradation persists in real-world clinical documentation

To understand diagnostic safety of medical AI systems in different languages, we conducted a prospective study in a real clinical setting. Fifty real ophthalmology dialogues produce d 150 AI-generated records. Physician correction was near-universal. Overall, 148 of 150 records (98·7%) required at least one field-level edit. The modification score was 53·6%, equal to about 3·2 corrected fields per record. This finding is central. It shows that multilingual degradation was not only a benchmark signal. It appeared as daily physician correction work. No pipeline was suitable for unsupervised documentation. Even the best pathway still required extensive human review.

The correction burden differs by language pathway. English-pivot had the lowest modificati on score (50·7%; 152/300 field slots). Chinese-direct was similar (51·0%; 153/300). Thai -pivot had the highest burden (59·0%; 177/300). Thai-pivot required more edited fields than Chinese-direct (p=0·004) and English-pivot (p=0·003). Chinese-direct and English-pivot were similar (p=0·686).

Edits were concentrated on chief complaints and history of present illness. These are clinically dense fields. Sampled edit-type analysis showed that most edits were substantive clinical corrections, not surface normalisation. Field-level patterns are detailed in Appendix 8. Past medical history and allergy history also showed important inter-pipeline differences, especially for Thai-pivot records. The prospective study therefore links the benchmark findings to real clinical speech.

These findings extend the benchmark results: multilingual degradation persisted in authentic clinical speech; routing through English introduced minimal overhead relative to direct Chinese extraction; and routing through Thai incurred a clinically meaningful penalty (+ 8·3 pp vs English-pivot, +8·0 pp vs Chinese-direct). The near-universal edit rate confirms that human physician oversight remained necessary across all pipelines.

### Benchmarks show language-dependent clinical degradation

GPT-series models showed a broad resource-ordered decline. On HealthBench, GPT-5.2 scores fell from 0·3743 in English to 0·3180 in Persian. In glaucoma classification, GPT-4o three-type accuracy fell from 50·7% in English to 32·7% in Persian. In ICU extraction, actionable-parameter recall fell from 100·0% in English to 48·5% in Persian. These drops affected clinical content, which were not only language-style differences. Qwen3 showed a different profile, with smaller or absent decline in some glaucoma settings. Full results are in Appendix 4 and eTable 1.

### Errors concentrate in multimodal and safety-critical tasks

The largest risks appeared when language was combined with images, structured clinical data, or urgent clinical subgroups. Aggregate accuracy often hides the problem. The common pattern was not random noise. It was a selective loss of the findings and values that clinicians most need for safe decisions.^22^

In OphthalmologyEHR-Glaucoma, adding bilateral fundus images amplified the language gap. G PT-4o three-type accuracy fell most in Persian. Chinese, Malay, and Thai also declined. Angle-closure glaucoma recall fell in several non-English settings. This matters because angle-closure disease is time-critical. High-risk glaucoma recall showed smaller degradation, which means exact diagnostic accuracy and safety recall captured different error types. In some low-resource settings the model preserved a broad high-risk label but distorted the specific disease subtype. Back-translation restored accuracy to 49·3–53·3% and improved angle-closure recall.^23^

In MIMIC-CXR, overall accuracy stayed near the English baseline, but macro-F1 fell from 0·2938 to 0·2149–0·2424. This pattern is clinically important. Reports could look glob ally acceptable while omitting pathological findings. The key risk was false-negative bias. A report that misses pneumothorax or consolidation can still sound coherent, but it fails the clinical task. Missed-critical-pathology results, patient-level any-critical-miss rates, and single-hazard recall are in Appendix 5.2.

In MIMIC-III, multilingual degradation targeted actionable physiology. GPT-4o actionable-parameter recall fell to 74·8% in Thai and 48·5% in Persian. Patient-level any-miss rates were 91·7% and 98·0%. Coarser variables, such as age and sex, were more stable. Continuous values that determine extubation safety were more vulnerable, including PaO2/FiO2, PEE P, vasopressor support, and acidaemia. The outputs were often fluent and showed acceptable apparent accuracy. This was partly because the task required only a binary yes/no decision with supporting reasoning. However, many responses were grounded in incorrect extraction of physiological information. Such extraction errors were more fundamental than reasoning errors and were easier to identify during evaluation.

### Safety-critical subgroups show larger gaps

Subgroup analysis showed that errors were not random. HealthBench emergency-referral quest ions had larger gaps than the overall task. In MIMIC-CXR, critical-pathology F1 declined more than routine-pathology F1. In glaucoma, angle-closure recall declined more than broader high-risk recall. In MIMIC-III, actionable abnormal parameters were more vulnerable than simple categorical fields. In the prospective study, initial visits required more editing than follow-ups. Optometry cases required more editing than general eye disease.

In HealthBench, emergency-referral scenarios showed the largest language gaps, with degradation 2·2–2·4 times greater than overall task-level decline. In MIMIC-CXR, the accuracy –F1 dissociation was driven by critical-pathology findings: critical-subset F1 fell from 30·8% in English to 20·0% in Chinese (–10·8 pp), whereas routine-subset F1 declined only from 27·9% to 23·0% (–4·9 pp). In OphthalmologyEHR-Glaucoma, angle-closure glaucoma recall, the most time-critical subtype, declined from 79·3% (English) to 66·9% (Thai), a larger drop than the broader high-risk group (worst: Malay 85·2% vs English 96·6%). In MIMIC-III, degradation was severity-weighted: coarser variables (age, sex, vital-sign stability) remained intact, but continuous parameters determining extubation safety (PaO_2_/FiO_2_, PEEP, vasopressor support, acidaemia) were most vulnerable to attenuation, substitution, o r smoothing. In the prospective study, initial visits required more editing than follow-up s (56·3% vs 52·5%), medium-complexity records had the highest burden (63·9%), and optometry cases (63·0%) exceeded general eye disease (53·0%). This pattern creates a compounded equity risk: patients in lower-resource languages face both more frequent and more consequential errors. (Subgroup definitions in Appendix 3; full stratified results in Appendix 10.)

### Confidence and reasoning signals do not provide a safety filter

Windowed perplexity confidence was unreliable for clinical triage. Across diagnostic task s, AUROC ranged from 0·41 to 0·66, mostly near chance. ECE ranged from 0·10 to 0·53. Correct and incorrect outputs received almost the same confidence. The Persian glaucoma case was especially concerning. The model assigned high confidence despite only 32·7% accuracy. This is more dangerous than visible uncertainty. Translation pivot reduced ECE in some settings, but it did not make perplexity confidence a validated safety signal.

Clinicians cannot rely on perplexity-based confidence to triage outputs. A PPL-based “auto -approve” threshold would be ineffective and potentially dangerous across all languages. Whether semantic entropy, self-consistency, or prompted self-evaluation offer better multilingual medical calibration remains an open question. More broadly, AUROC and ECE quantify different properties: a model may weakly rank correct above incorrect outputs while assigning systematically inflated confidence. AUROC alone is insufficient for clinical reliability; confidence analysis should be interpreted jointly with prespecified clinical risk measures.

### Reasoning instability clusters in the middle of the chain

Token-level analysis was available only for GPT-4o. Cross-lingual uncertainty concentrated around 40–70% of the reasoning trajectory. This is the stage where the model integrates evidence and commits to an interpretation. Back-translation reduced several of these gaps.

Logic-break rates rose as language resource level decreased. In MIMIC-III, they reached 8 6·2% in Persian and 84·3% in Thai, versus 40·8% in English. Mutation counts were higher in incorrect outputs than in correct outputs. Case examples in Appendix 11 show that these failures changed clinical content, not only scores. They changed infant-feeding advice, missed pneumothorax, distorted glaucoma severity, and smoothed ICU physiology into falsely reassuring narratives.

### Back-translation reduces degradation without retraining

Back-translation pivot was the most consistent training-free mitigation. It improved GPT-5.2 HealthBench scores in all non-English languages. It restored GPT-4o glaucoma accuracy to 49·3–53·3%. It restored MIMIC-CXR F1 to 0·2978–0·3432. It also restored MIMIC-III actionable recall to 100·0% for GPT-4o and 99·3–99·9% for Qwen3. The largest gains app eared in endpoints with direct safety relevance, including urgent HealthBench scenarios, critical-pathology F1, angle-closure recall, and actionable ICU variables.^20^

Back-translation also reduced process-level instability. Logic-break gaps narrowed, and EC E improved in several settings. English chain-of-thought was less consistent and sometimes unhelpful. These findings support back-translation as a practical mitigation. They do not make it a safety guarantee. Pivot outputs sometimes converged on simplified English templates and could still omit findings. The prospective study shows why translation strategies still need clinical validation before deployment.

## Discussion

### Main interpretation

The central finding is simple. Language changed the safety of medical AI outputs. The failures were clinically directed. They included missed radiology findings, missed ICU parameters, and glaucoma misclassification. English-only validation would not detect these error s. The prospective study confirmed that this risk appears in real clinical documentation. This is the main strength of the manuscript. It moves the problem from benchmark performance into clinical workflow.

### Model-specific safety profiles

The pattern was not identical across model families. GPT-4o/GPT-5.2 showed stronger resource-ordered degradation. Qwen3 showed smaller decline in some glaucoma settings. This may reflect differences in multilingual pretraining, tokenisation, visual encoders, alignment, or parameter scale. The mechanism cannot be proven from these experiments. The practical message is still clear. A safety result from one model should not be transferred to another model without language-specific testing.

Qwen3’s advantage in certain non-English glaucoma settings likely reflects alignment between native-language reasoning and diagnostic label semantics (PACS, PAC, PACG, APAC, secondary glaucoma), combined with shorter reasoning traces that reduce over-elaboration. Broader multilingual pretraining (∼36 trillion tokens across 119 languages) is a plausible but unproven contributor. GPT-4o and Qwen3 also differ in architecture, tokenisation, visual encoders, alignment training, and parameter scale, any of which could affect multilingual safety.

Two implications follow. First, multilingual degradation is especially prominent in English-dominant training regimes but not necessarily universal across all models. Second, multi lingual safety profiles cannot be generalised across model families; deployment decisions require evaluation of the specific model and workflow.

### Clinical equity risk

The most important errors occurred where clinical stakes were high. Critical-pathology rec all, angle-closure glaucoma recall, and actionable ICU extraction were more vulnerable than simpler outputs. Patients in lower-resource-language settings may therefore face both more frequent errors and more dangerous errors. This makes multilingual AI safety a health equity issue, not only a technical benchmark issue. In settings where language diversity is highest, safeguards for AI monitoring are often fewest; a model validated only in English should not be assumed safe in other languages.^21^

### Prospective study as the key validation

The prospective clinical study is the strongest evidence in this manuscript. It tested rea l outpatient speech, not a curated benchmark. It showed that nearly every AI-generated record needed physician correction. Most sampled edits were substantive clinical changes. The Thai-pivot pathway produced a higher correction burden than the English-pivot and Chinese-direct pathways. This result supports the need for physician oversight across all evaluated language pathways.

The prospective study also gives a practical endpoint. The required modification score is easy to audit. It shows how much physician work is needed to make AI documentation usable. It can be stratified by field, visit type, disease type, and language pathway. This endpoint can support future multilingual quality monitoring, especially after multi-site validation.

### Confidence is not a clinical safety filter

Perplexity-based confidence did not separate correct from incorrect outputs. This was true across languages and tasks. A threshold based on this signal would miss unsafe outputs and may create false reassurance. AUROC and ECE should also be interpreted together. A model m ay rank outputs weakly while still being overconfident. Better uncertainty methods require prospective testing against clinical endpoints.^25^

### Reasoning instability and mitigation

Token-level analysis suggests that GPT-4o multilingual degradation reflects instability during reasoning execution. The key signal appeared in the middle of the chain. Back-translation moved inference into a more stable linguistic space and improved performance.

This mechanism remains a hypothesis. Back-translation may also help by standardising terminology or improving prompt-model alignment. Token-level traces were unavailable for GPT-5. 2 and Qwen3, so process-level claims should not be generalised across models.

### Strengths and limitations

This study combined four clinical tasks, five languages, multiple inference paradigms, token-level analysis, cross-model comparison, and a prospective clinical study. It also has limitations. The prospective study was single-centre and ophthalmology-specific. It used Chinese source dialogues and one physician reviewer. The language set did not include Arabic, Hindi, Swahili, or many other clinically important languages. Token-level analysis was limited to GPT-4o. Benchmark sample sizes were modest. Future work should test more sites, more languages, more reviewers, and more clinical workflows.

### Policy implications

Clinical AI governance should require language-specific safety auditing when systems are deployed in multilingual settings. Audits should include clinical risk endpoints, not only aggregate accuracy. Human oversight should remain in AI-generated documentation. This recommendation is directly supported by the prospective study. Modification score can provide a practical monitoring metric, but it needs multi-site validation. Developers should also report per-language safety profiles. The open-source toolkit released with this study provides one way to start this auditing work. Extended recommendations are in Appendix 12.^24^

## Data Availability

All data produced in the present study are available upon reasonable request to the authors

## Contributors

YQW, HYH, and RPZ contributed equally. YQW and HYH designed the study. YQW, RPZ, and YYL wrote and ran the code, performed benchmark evaluations, and summarized results. YQW performed the statistical analysis and drafted the manuscript. PP, AK, YZ, and SPC assessed translated materials. MQP assessed the real-world clinical relevance of the findings. ZPL, KZ, YCT, HD, CYC, and JY advised on experimental design and manuscript preparation. DBL supervised the study and is the corresponding author. All authors reviewed and approved the final manuscript.

## Data sharing statement

Analysis code, prompt templates, and figure-generation scripts are available at https://github.com/wang-ava/languagegap. The open-source safety auditing toolkit is available at https://doctor-language-bridge.onrender.com/. HealthBench data are available through OpenAI. MIMIC-CXR and MIMIC-III data are available through PhysioNet under credentialled data use agreements. The institutional ophthalmology EHR dataset and de-identified prospective study data are available from the corresponding author upon reasonable request, subject to institutional data governance requirements.

