## Appendix for "Language-dependent diagnostic safety of medical AI systems: a cross-lingual benchmarking and prospective clinical study"

### Supplementary Appendix

#### Appendix.1 Study Design, Model Selection, Inference-Paradigm Applicability, and Translation Checks

This appendix provides methodological detail that is intentionally abbreviated in the manuscript: model selection, task-specific inference-paradigm applicability, translation controls, and the scope of token-level analysis.

The benchmark programme linked translated clinical inputs to four task formats: HealthBench conversational medical question answering, OphthalmologyEHR-Glaucoma classification, MIMIC-CXR radiology report generation, and MIMIC-III re-intubation risk prediction. English was the source and reference condition; Chinese, Malay, Thai, and Persian were chosen to span high- to low-resource language representation.

The prospective component was kept separate from the benchmark programme and used real ophthalmology outpatient dialogues to test whether benchmark failure modes appeared as physician correction burden in clinical documentation.

Model selection reflected task-specific constraints. GPT-5.2 was used for HealthBench because GPT-4o produced near-zero non-English scores under HealthBench's strict rubric, while GPT-4o was retained for OphthalmologyEHR-Glaucoma, MIMIC-CXR, and MIMIC-III. Qwen3 VL 235B A22B Thinking was included as a multilingual reference model because its broader multilingual pretraining provides a useful comparison with the English-dominant GPT series.

Three inference paradigms were tested. Native-language reasoning used the target language for question, reasoning, and answer. English chain-of-thought asked the model to reason in English and answer in the target language. Back-translation pivot translated the target-language input into English before English-language reasoning and answer generation.

Setting 2 was not applied to HealthBench or MIMIC-CXR because their native formats do not require explicit structured chain-of-thought outputs. HealthBench evaluates free-form conversational answers, whereas MIMIC-CXR requires direct radiology report generation from images, with reports naturally encoding observations, findings, and clinical impressions. Requiring explicit English intermediate reasoning would alter the response format and introduce confounds incompatible with the original benchmark criteria. Therefore, Setting 2 was limited to OphthalmologyEHR-Glaucoma and MIMIC-III, where structured JSON or reasoning-step outputs were inherent to the task design.

Token-level behavioural analyses were restricted to GPT-4o because per-token log-probability parameters are not exposed by OpenAI's reasoning model series, including GPT-5.2, or by Qwen3 VL 235B A22B Thinking. Therefore, position-wise perplexity curves, logic-break rates, mutation counts, and related mechanistic interpretations apply specifically to GPT-4o and should not be generalised directly to GPT-5.2 or Qwen3.

Translation prompts preserved medical terminology, numerical values, measurement units, abbreviations, and negation. Clinicians rated forward translations for clinical accuracy, clinical safety, and usability; Chinese and Malay reached near-ceiling ratings, Thai was lower but remained clinically usable, and Persian remained high but below ceiling.

Back-translation metrics showed strong preservation of clinically important content. Number F1 and Number+Unit F1 exceeded 0·95 across languages, and negation F1 ranged from 0·908 to 0·942. Surface lexical metrics were lower because valid paraphrase changed wording, supporting the interpretation that major translation errors were not the primary cause of performance gaps.

Subgroup, calibration, and token-level analyses were handled as separate analytical modules. Subgroup definitions are given in Appendix 3, calibration definitions in Appendix 2, and token-level methods in Appendix 7.

#### Appendix.2 Confidence Calibration Metrics and Windowed Perplexity Definition

Standard full-sequence perplexity was not used as the primary confidence proxy because gen erated outputs often begin with template phrasing and end with formatting tokens. Windowed PPL was therefore defined over the middle 60% of generated tokens, corresponding to positi ons 20%–80%, to retain the core reasoning segment while attenuating template and formatti ng effects. Confidence was defined as confidence20–80 = 1/PPL20–80.

Four calibration metrics were computed. AUROC measured whether confidence could rank corre ct outputs above incorrect outputs, treating correctness as the positive class and confide nce20–80 as the score variable. Expected calibration error was computed by partitioning s amples into equal-frequency confidence bins and calculating the weighted absolute differen ce between mean confidence and empirical accuracy within each bin. The confidence separati on gap was defined as the mean confidence among correct outputs minus the mean confidence among incorrect outputs. The top-bottom accuracy gap was defined as the accuracy of the hi ghest-confidence quartile minus the accuracy of the lowest-confidence quartile.

Higher AUROC, confidence separation gap, and top-bottom accuracy gap indicate better calib ration behaviour, whereas lower ECE indicates better calibration. These four metrics were used jointly to capture ranking ability, calibration quality, mean confidence separation, and the practical utility of confidence for prioritising more reliable outputs.

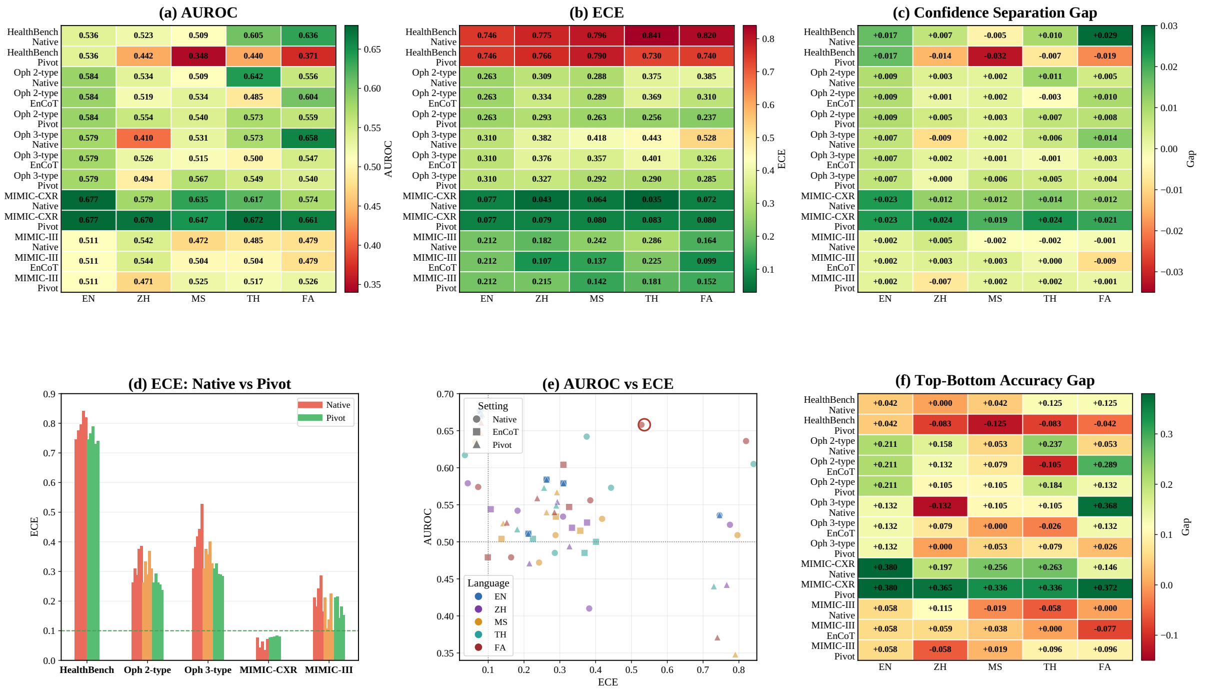

***eFigure 1. Calibration performance of GPT-4o across languages, datasets, and inference se ttings.*** *Panels show (a) AUROC, (b) expected calibration error (ECE), (c) confidence separa tion gap, (d) ECE comparison between native-language inference and back-translation pivot, (e) joint AUROC–ECE distribution, and (f) top-bottom accuracy gap. Confidence was defined as inverse perplexity over the middle 20%–80% of generated tokens. In panel (e), marker s hape denotes inference setting, marker color denotes language, and bubble size denotes the absolute confidence separation gap. Higher AUROC, confidence separation gap, and top-botto m accuracy gap indicate better calibration, whereas lower ECE indicates better calibratio n. EN, English; ZH, Chinese; MS, Malay; TH, Thai; FA, Persian.*

#### Appendix.3 Subgroup Definitions for Safety and Fairness Analyses

For HealthBench text QA, benchmark theme annotations were used to define a high-acuity subgroup consisting of emergency-referral questions and a high-complexity subgroup consisting of multi-step clinical reasoning questions. Overall score degradation across all samples was compared with within-subgroup degradation for each language.

For MIMIC-CXR, the 14 CheXpert labels were stratified into an urgent-care subset and a routine subset. The urgent-care subset included pneumothorax, pneumonia, oedema, consolidation, pleural effusion, cardiomegaly, and enlarged cardiomediastinum. The routine subset included atelectasis, lung lesion, lung opacity, fracture, no finding, support devices, and pleural other. Macro-averaged F1 was computed within each subset for each language and inference method. Differential degradation was quantified as $\Delta F1(CRITICAL) - \Delta F1(ROUTINE)$ relative to the English baseline.

For OphthalmologyEHR-Glaucoma, disease-subtype stratification compared degradation in the broader high-risk glaucoma group with degradation in the angle-closure subgroup, the most time-critical subtype group. For MIMIC-III, degradation on actionable parameters, defined as values outside the normal range, was compared with overall parameter extraction performance.

For the prospective study, modification scores were stratified by disease type, visit type, and record quality level, comparing eye disease versus optometry, initial versus follow-up visits, and high-, medium-, versus low-quality records. Patient-level demographic stratification by age and gender was not feasible because these variables were not available in the benchmark evaluation pipeline.

#### Appendix.4 Full Cross-Language Benchmark Results and Safety-Specific Error Patterns

##### Appendix.4.1 Cross-Language Degradation and Safety-Specific Endpoints

Across native-language inference, GPT-series models showed a broadly resource-ordered degradation pattern. Because GPT-5.2 was evaluated on HealthBench and GPT-4o on the remaining datasets, these findings should be interpreted as family-level descriptive trends rather than controlled within-model comparisons. MIMIC-III Stage 2 binary accuracy did not follow the same pattern and in some cases inverted it, likely because binary accuracy was near chance and therefore less informative than Stage 1 safety-relevant extraction metrics. Qwen3 VL 235B A22B Thinking showed a partially different multilingual profile, including attenuated degradation in some multimodal settings.

Across datasets, safety-specific endpoints were interpreted alongside aggregate performance because multilingual failures were not always captured by overall accuracy. In HealthBench, degradation was interpreted in relation to missed urgent escalation, omitted safety-critical counselling, and unsafe recommendations. In OphthalmologyEHR-Glaucoma, safety interpretation focused on high-risk glaucoma and angle-closure glaucoma recall. In MIMIC-CXR, the main safety concern was missed pathological findings despite preserved report-level accuracy. In MIMIC-III, the main safety endpoint was actionable-parameter recall, defined as extraction of abnormal values requiring potential clinical attention.

| Dataset / Metric | English | Chinese | Malay | Thai | Persian | English | Chinese | Malay | Thai | Persian |
|---|---|---|---|---|---|---|---|---|---|---|
| HealthBench text QA (, n=100) | | | | | | | | | | |
| **model** | **GPT-5.2** | | | | | **Qwen3 VL 235B A22B Thinking** | | | | |
| English criterion score Native Reasoning | 0•3743± 3•11% | 0•3658▼± 3•421% | 0•3498▼± 3•59% | 0•3327▼± 3•59% | 0•3180▼± 3•48% | 0•3050± 2•98% | 0•2030▼± 3•30% | 0•1449▼± 3•45% | 0•1627▼± 3•38% | 0•1313▼± 3•52% |
| Specific criterion score Native Reasoning | 0•3743± 3•11% | 0•3544▼± 3•521% | 0•3459▼± 3•35% | 0•3008▼± 3•47% | 0•3311▼± 3•43% | 0•3050± 2•98% | 0•2230▼± 3•41% | 0•2375▼± 3•28% | 0•2625▼± 3•55% | 0•1681▼± 3•60% |
| English CoT | HealthBench does not require structured CoT output | | | | | | | | | |
| Back-translation | 0•3743± 3•11% | 0•4240†± 3•20% | 0•4237†± 3•74% | 0•4196†± 3•04% | 0•4254†± 3•03% | 0•3050± 2•98% | 0•2663± 3•15% | 0•3051†± 3•62% | 0•2476± 3•22% | 0•3203†± 3•10% |
| **OphthalmologyEHR-Glaucoma 2-type / text-only (n=150 patients)** | | | | | | | | | | |
| **model** | **GPT-4o** | | | | | **Qwen3 VL 235B A22B Thinking** | | | | |
| Accuracy Native Reasoning | 53•3%± 7•88% | 52•0%▼± 7•89% | 53•3%± 7•88% | 45•3%▼± 7•90% | 48•7%▼± 7•87% | 50•0%± 7•90% | 58•0% ± 7•80% | 55•3%± 7•86% | 56•0%± 7•85% | 52•0%± 7•89% |
| Accuracy English CoT | 53•3%± 7•88% | 48•0%± 7•89% | 52•0%± 7•89% | 49•3% ± 7•87% | 45•3%± 7•90% | 50•0%± 7•90% | 52•7%± 7•89% | 54•0%± 7•88% | 52•7%± 7•89% | 54•0%± 7•88% |
| Accuracy Back-translation | 53•3%± 7•88% | 50•7%± 7•90% | 53•3%†± 7•88% | 56•0%†± 7•88% | 54•0%†± 7•85% | 50•0%± 7•90% | 52•7%†± 7•89% | 54•7%†± 7•87% | 52•7%†± 7•89% | 54•7%†± 7•87% |
| **MIMIC-CXR radiology report generation (n=150 patients)** | | | | | | | | | | |
| **model** | **GPT-4o** | | | | | **Qwen3 VL 235B A22B Thinking** | | | | |
| Accuracy Native Reasoning | 0•7955± 2•85% | 0•7775▼± 3•03% | 0•7611▼±3•58% | 0•7861▼± 3•10% | 0•7838▼± 3•24% | 0•7805± 3•34% | 0•7355▼± 3•64% | 0•7310▼± 3•83% | 0•7212▼± 4•03% | 0•7282▼± 3•77% |
| Accuracy Back-translation | 0•7955± 2•85% | 0•7978†± 2•88% | 0•7915± 2•94% | 0•8019†± 2•76% | 0•7967†± 2•87% | 0•7805± 3•34% | 0•7793± 3•37% | 0•7838± 3•33% | 0•7839± 3•33% | 0•7878± 3•22% |
| F1 Native Reasoning | 0•2938± 5•86% | 0•2149▼±5•10% | 0•2296▼± 5•56% | 0•2424▼± 5•89% | 0•2187▼± 5•51% | 0•3369± 5•29% | 0•3218▼± 5•38% | 0•3069▼± 5•93% | 0•2960▼± 5•71% | 0•2973▼± 5•81% |
| F1 Back-translation | 0•2938± 5•86% | 0•3046†± 5•80% | 0•2978†± 5•77% | 0•3432†± 5•28% | 0•3092†±5•86% | 0•3369± 5•29% | 0•3398†± 5•78% | 0•3321± 5•69% | 0•3712†± 5•35% | 0•3557†± 5•29% |
| English CoT | MIMIC-CXR does not require structured CoT output | | | | | | | | | |
| **OphthalmologyEHR-Glaucoma 3-type / text + fundus images (n=150 patients)** | | | | | | | | | | |
| **model** | **GPT-4o** | | | | | **Qwen3 VL 235B A22B Thinking** | | | | |
| Accuracy Native Reasoning | 50•7%± 7•90% | 46•7%▼± 7•88% | 42•7%▼± 7•82% | 43•3%▼± 7•83% | 32•7%‡▼± 7•42% | 53•3%± 7•88% | 53•3%± 7•88% | 53•3%± 7•88% | 52•7%▼± 7•89% | 54•0%± 7•88% |
| Accuracy English CoT | 50•7%± 7•90% | 45•3%± 7•87% | 46•7%± 7•88% | 44•0%± 7•85% | 50•0%± 7•90% | 53•3%± 7•88% | 55•3%± 7•86% | 54•0%± 7•88% | 60•0%± 7•75% | 52•0%± 7•89% |
| Accuracy Back-translation | 50•7%± 7•90% | 49•3% ± 7•90% | 52•7%†± 7•89% | 52•7%†± 7•89% | 53•3%†± 7•88% | 53•3%± 7•88% | 53•3%± 7•88% | 53•3%± 7•88% | 52•7%± 7•89% | 57•3%†± 7•82% |
| **MIMIC-III ICU re-intubation prediction, Stage 2 (n=195 patients)** | | | | | | | | | | |
| **model** | **GPT-4o** | | | | | **Qwen3 VL 235B A22B Thinking** | | | | |
| Accuracy Native Reasoning | 47•8%± 6•77% | 45•4%▼± 6•75% | 49•3%± 6•78% | 47•3%▼± 6•77% | 52•7%± 6•77% | 50•7%± 6•78% | 50•7%± 6•78% | 53•2%± 6•77% | 45•6%▼ ± 6•92% | 50•0%▼± 6•86% |
| Accuracy English CoT | 47•8%± 6•77% | 52•0%± 6•79% | 52•7%± 6•77% | 44•4%± 6•74% | 50•2%± 6•78% | 50•7%± 6•78% | 52•2%± 6•77% | 54•1%± 6•76% | 44•9%± 6•75% | 54•1%± 6•76% |
| Accuracy Back-translation | 47•8%± 6•77% | 47•8%††± 6•77% | 55•1%††± 6•75% | 50•7%††± 6•78% | 53•7%††± 6•76% | 50•7%± 6•78% | 53•2%††± 6•77% | 53•2%††± 6•77% | 51•7%††± 6•77% | 52•7%††± 6•78% |

***Table 1. Cross-lingual performance across four medical benchmarks and five languages under three inference paradigms.*** *Results are reported under native reasoning, English CoT constraint, and back-trans ation pivot. ▼ indicates performance below the English native baseline; † indicates performance equal to or above the English baseline after back-translation. ‡ highlights Persian three-type glaucoma a curacy of 32•7%. †† indicates that MIMIC-III Stage 2 binary accuracy should be interpreted with Stage 1 extraction metrics. CoT = chain of thought; F1 = harmonic mean of precision and recall. Values are eported as point estimate ± half-width of bootstrap 95% CI (10 000 resamples). For example, 0•3743 ± 3•11% corresponds to a 95% CI of approximately 0•363–0•386.*

##### Appendix.4.2 HealthBench Extended Results

HealthBench showed a monotonic GPT-5.2 decline under native-language inference from English to Persian: 0·3743 in English, 0·3658 in Chinese, 0·3498 in Malay, 0·3327 in Thai, and 0·3180 in Persian. Qwen3 showed a lower English baseline and larger cross-lingual gap, declining from 0·3050 in English to 0·1313 in Persian. A criterion-language effect was observed for GPT-5.2, including Thai scoring 0·3327 under English rubric criteria versus 0·3008 under Thai-language criteria, but deficits persisted after accounting for rubric language. Back-translation improved GPT-5.2 HealthBench scores across all non-English languages to 0·4196–0·4254 and improved Qwen3 performance in several languages, including Malay and Persian.

##### Appendix.4.3 MIMIC-III Extended Results

Under GPT-4o native-language inference, actionable-parameter recall was 100·0% in English but fell to 74·8% in Thai and 48·5% in Persian. At the patient level, 91·7% of Thai-language patients and 98·0% of Persian-language patients had at least one missed actionable parameter, compared with 0·0% in English. Re-intubation recall also declined from 49·4% in English to 39·3% in Persian. Qwen3 showed a similar low-resource failure pattern, with actionable-parameter recall falling to 62·9% in Thai and 61·1% in Persian and patient-level any-miss rates of 99·0% and 99·5%, respectively. English-CoT did not improve these rates, whereas translation pivot restored actionable-parameter recall to 100·0% across all languages for GPT-4o and 99·3–99·9% for Qwen3.

The clinically important failure mode in MIMIC-III was missed actionable physiology. Values affecting extubation safety, including PaO2/FiO2, PEEP, vasopressor support, and acidaemia, were more vulnerable than simpler fields. Outputs could remain fluent while giving falsely reassuring low-risk assessments.

#### Appendix.5 Non-Text and Multimodal Safety Degradation

##### Appendix.5.1 OphthalmologyEHR-Glaucoma Extended Results

In the two-type setting, GPT-4o accuracy was close to the English baseline of 53·3% for Chinese and Malay, while Thai and Persian declined to 45·3% and 48·7%. Back-translation recovered performance more reliably than English-CoT, with Thai and Persian reaching 56·0% and 54·0%. Qwen3 showed no resource-ordered degradation in this text-only setting.

Safety recall was computed at the eye level using clinically grouped risk categories rather than exact diagnostic-code match. High-risk glaucoma recall, defined as PACG, APAC, POAG, and secondary glaucoma, showed smaller but still clinically relevant degradation; the worst GPT-4o three-type case was Malay at 85·2% versus 96·6% in English. Translation pivot restored angle-closure recall to near-baseline levels across languages, ranging from 72·7% to 80·2% compared with 79·3% in English. Qwen3 showed smaller safety degradation, with the worst-case angle-closure recall being 81·8% for Thai under English-CoT in the two-type setting, compared with 85·1% in English.

Safety recall and exact accuracy measure different error types. Some non-English settings showed higher high-risk glaucoma recall than English, such as GPT-4o two-type Persian at 98·3% versus 94·3% in English. This does not necessarily indicate better diagnostic performance, because misclassification within the high-risk group, such as PACG classified as APAC, preserves high-risk recall while reducing exact accuracy. In low-resource languages, uncertainty may lead the model to default to broader high-risk categories, increasing recall but also increasing false positives and unnecessary referrals. The angle-closure subgro up, which requires more precise subtype discrimination, showed more consistent degradation and is therefore more informative for time-critical safety risk.

The text-only two-type glaucoma setting showed smaller gaps than the text-plus-fundus three-type setting. In the three-type GPT-4o setting, Persian accuracy fell to 32·7% compared with 50·7% in English, while back-translation restored performance to near-baseline levels.

##### Appendix.5.2 MIMIC-CXR Extended Results

Under GPT-4o native-language inference, accuracy remained near the English baseline of 0·7955 across non-English languages, ranging from 0·7611 to 0·7861. However, macro-F1 declined from 0·2938 in English to 0·2149–0·2424 across non-English languages. Qwen3 showed smaller F1 declines, from an English baseline of 0·3369 to 0·2960–0·3218, and back-translation improved Qwen3 F1 to 0·3321–0·3712, although Malay remained slightly below the English baseline.

Safety-specific endpoints included missed-critical-pathology rate, patient-level any-critical-miss rate, and single-hazard recall for urgent findings such as pneumothorax. These were interpreted separately from overall accuracy and macro-F1 because a report can be linguistically plausible and globally accurate while omitting the finding most relevant to patient management.

MIMIC-CXR showed an accuracy-F1 dissociation. Overall accuracy stayed near the English baseline, but macro-F1 fell in non-English languages, raising concern for false-negative bias. A report can appear acceptable while omitting a clinically critical finding such as pneumothorax.

#### Appendix.6 Back-Translation as a Cross-Task Mitigation Pattern

Back-translation was analysed as a cross-task mitigation pattern rather than as a separate model. It most consistently improved endpoints that were clinically safety-relevant: urgent HealthBench scenarios, multimodal glaucoma accuracy, MIMIC-CXR macro-F1, and MIMIC-III actionable-parameter recall. Qwen3 also improved under back-translation in several tasks, particularly MIMIC-CXR and MIMIC-III.

English-CoT was less consistent. It was not applied to HealthBench or MIMIC-CXR because their native task formats did not require explicit structured reasoning, and in OphthalmologyEHR-Glaucoma and MIMIC-III it produced task-dependent effects that did not reliably improve safety-specific endpoints. This supports the main-text interpretation that translation before reasoning was more robust than imposing English intermediate reasoning while preserving non-English final outputs.

The interpretation is therefore pragmatic: translation before reasoning may move the model into a more stable linguistic operating space, but it does not remove the need for language- and workflow-specific clinical validation.

#### Appendix.7 Token-Level Behavioural Analysis and Reasoning Instability

##### Appendix.7.1 Token-Level Behavioural Analysis Methodology

Token-level log-probability traces were recorded for each generated token from GPT-4o. Three complementary metrics were computed to characterise reasoning instability.

Position-wise perplexity curves quantified stage-dependent uncertainty accumulation by computing per-token perplexity, $\exp(-\log p)$, normalising token position to a 0–1 scale, and averaging values across 20 equal-width bins.

Logic-break rate was defined as the proportion of samples containing at least one token for which the adjacent log-probability drop exceeded $\tau\_drop = 1 \cdot 5$ and either the standardised drop exceeded $\tau\_z = 2 \cdot 0$ or the absolute log-probability was less than $\tau\_abs = -2 \cdot 5$.

Mutation count quantified trajectory volatility as the number of log-probability jumps with absolute magnitude of at least 2·0; distributions were visualised with violin plots stratified by output quality. Stage localisation used 10 normalised position buckets with mean log-probability per bucket, displayed as cross-lingual heatmaps. These three metrics jointly captured stage-dependent uncertainty accumulation, discrete reasoning discontinuities, and trajectory volatility along the chain-of-thought.

Confidence calibration methodology and the four metrics (AUROC, ECE, confidence separation gap, top-bottom accuracy gap) are defined in Appendix 2 and not repeated here.

##### Appendix.7.2 Reasoning Instability Extended Results

Across text-only tasks, position-wise perplexity curves and mean log-probability heatmaps showed that cross-lingual uncertainty concentrated in the mid-chain inference window, approximately 40–70% of the normalised trajectory, with a smaller rise during answer consolidation. Back-translation pivot attenuated these mid-chain gaps. Logic-break rates increased with decreasing language resource level and were reduced by back-translation. Mutation counts were wider and higher for incorrect than correct outputs, indicating trajectory volatility correlated with task-level failure. In vision-language and dashboard-based tasks, including OphthalmologyEHR-Glaucoma three-type, MIMIC-CXR, and MIMIC-III, instability extended to the junctions between visual grounding and linguistic explanation.

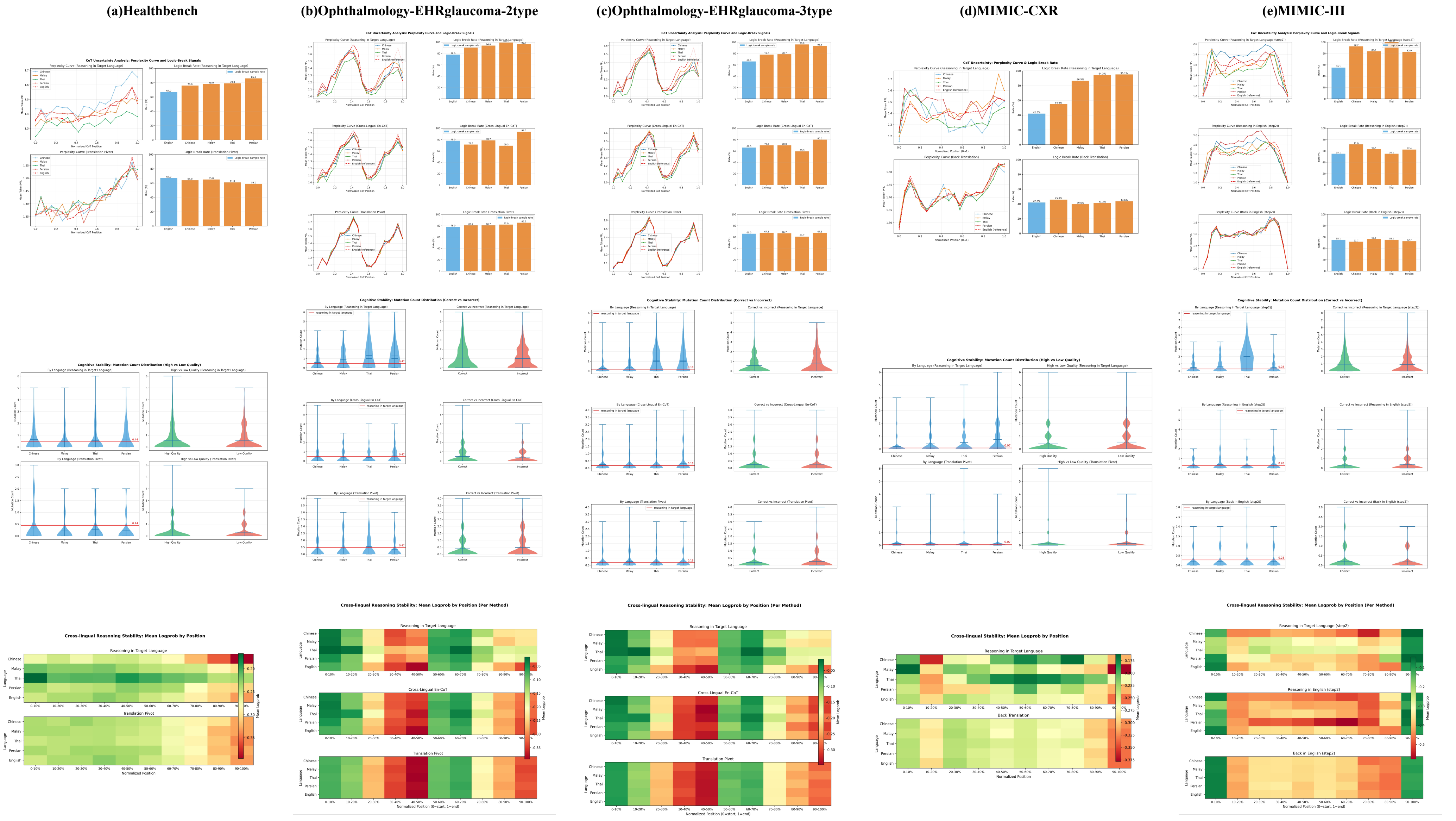

*eFigure 2. Token-level behavioural analysis of reasoning instability across tasks and applicable inference paradigms.* For each dataset, three panel groups are presented. (Top) Chain-of-thought uncertainty analysis: position-wise perplexity curves by language and logic-break rate bar plots, shown separately for native-language inference and back-translation pivot, with English chain-of-thought constraint shown only for OphthalmologyEHR-Glaucoma and MIMIC-III. (Middle) Cognitive stability: mutation count violin plots stratified by output quality. (Bottom) Cross-lingual reasoning stability: mean log-probability by normalised position heatmaps. Panels: (a) HealthBench; (b) OphthalmologyEHR-Glaucoma 2-type; (c) OphthalmologyEHR-Glaucoma 3-type; (d) MIMIC-CXR; (e) MIMIC-III. CoT =chain of thought.

#### Appendix.8 Prospective Study and Extended Analysis

##### Appendix.8.1 Prospective Study Details

The source corpus comprised 165 de-identified audio-transcribed clinician–patient dialogue records from a real-world ophthalmology outpatient clinic. Dialogues were quality-graded at source: 136 were classified as high-quality, defined as at least five dialogue turns with complete, legible content, and 29 as low-quality, defined as fewer than five turns or predominantly dialectal content. A stratified random sample of 50 dialogues was drawn proportionally from the quality strata. Dialogues were fully de-identified prior to use.

Each of the three pipelines (Chinese-direct, English-pivot, and Thai-pivot) produced a structured medical record with six standardised editable fields: chief complaint, history of present illness (HPI), past medical history, allergy history, family history, and personal history. Non-Chinese outputs from the English-pivot and Thai-pivot variants were subsequently translated into Chinese to enable unified physician review. The modification score for each pipeline was computed as modified field slots divided by the number of samples multiplied by six editable fields. Physician review was conducted using a structured interface enabling field-level annotation.

The overall required modification score was 53·6% (Figure 3A). Chinese-direct and English-pivot performed comparably (51·0% and 50·7%), whereas Thai-pivot required more modification (59·0%). At the record level, 148 of 150 records (98·7%) required at least one physician edit: 96% for Chinese-direct and 100% for both English-pivot and Thai-pivot (Figure 3B). Paired Wilcoxon signed-rank tests confirmed that Thai-pivot required more edited fields than Chinese-direct ($p=0·004$) or English-pivot ($p=0·003$), while Chinese-direct and English-pivot did not differ ($p=0·686$) (Figure 3F).

To characterise edit types, a random sample of 20 edited fields was drawn from each pipeline’s reviewed records for qualitative classification into one of four categories: clinical detail extraction, history/risk correction, uncertainty resolution, or surface normalisation.

The modification score is a workflow-quality endpoint rather than a direct safety-critical error rate, because each field-level edit is weighted equally. To separate workflow burden from clinical risk, sampled edits were classified as clinical detail extraction, history/risk correction, uncertainty resolution, or surface normalisation. Most sampled edits were substantive rather than surface-level (18/20 in Chinese-direct and 16/20 in English-pivot) (Figure 3E). Future studies should scale a graded edit taxonomy that distinguishes stylistic, clinically relevant, and safety-critical corrections.

##### Appendix.8.2 Field-Level Details

Field-level modification rates varied markedly across the six documentation fields (eFigure 3A, Figure 3C).

Personal history required no edits across any pipeline or quality stratum (0% modification for Chinese-direct, English-pivot, and Thai-pivot alike; 0% in both high-quality and low-quality dialogues). Empty-field analysis confirmed that personal history was almost entirely empty in both the source dialogues and AI outputs (source and AI both empty in close to 100% of records), indicating negligible extractable information rather than superior extraction performance.

Family history required uniformly few edits (8% modification across all three pipelines; 7% in high-quality and 11% in low-quality records). The completeness decomposition showed that approximately 50% of family history records had both source and AI fields empty, with most of the remaining records classified as non-empty and unchanged by the physician, and only a small proportion requiring modification.

Chief complaint and HPI were non-empty in nearly all records but required physician modification in the vast majority of cases. For chief complaint, modification rates were 88% (Chinese-direct), 98% (English-pivot), and 94% (Thai-pivot). For HPI, modification rates were 88% (Chinese-direct), 86% (English-pivot), and 98% (Thai-pivot). The completeness decomposition showed that both fields were dominated by the "non-empty field changed" category (approximately 93% and 90% respectively in the aggregate), with only thin slivers of unchanged content (approximately 7% and 10%).

Past medical history showed the largest inter-pipeline divergence: the Thai-pivot variant required modification in 92% of records, compared with 72% for Chinese-direct and 66% for English-pivot. The completeness decomposition showed approximately 77% of records required changes in aggregate, with the remaining approximately 23% classified as non-empty and unchanged.

Allergy history exhibited a distinctive pattern. The completeness decomposition revealed a notable proportion of records in which the source field was empty but the AI nonetheless generated content ("AI filled despite empty source"), alongside records with non-empty fields that were changed and a smaller grey (both empty) component. Modification rates diverged across pipelines: 62% for Thai-pivot, 50% for Chinese-direct, and 46% for English-pivot.

**Appendix.8.3 Edited-Field Distributions**

Most records required edits in at least two fields across all three pipelines (Figure 3D). The distribution of edited fields per record was categorised into three bands: 0–1 fields, 2–3 fields, and 4–6 fields. Approximately half of all records required edits in four or more fields, indicating widespread moderate correction rather than a small number of extreme failures. The Thai-pivot pipeline showed the largest proportion of records in the 4–6 edited fields band, consistent with its higher overall modification score (59·0%). Chinese-direct and English-pivot showed comparable distributions, with a somewhat larger share of records in the 2–3 band relative to Thai-pivot. Only a small minority of records across any pipeline required zero or one edit.

**Appendix.8.4 Dialogue Quality Effects**

Low-quality dialogues (n=27 generated records) had consistently higher field-level modification proportions than high-quality dialogues (n=123 generated records) across all non-trivial fields (eFigure 3B). The largest absolute difference appeared in allergy history (81% for low-quality vs 46% for high-quality, a 35-percentage-point gap). Past medical history also showed a notable increase (85% vs 75%, a 10-percentage-point gap). Chief complaint (96% vs 93%) and HPI (96% vs 89%) showed smaller but consistent increases. Personal history remained at 0% modification in both strata. Family history showed only a modest increase (11% vs 7%). These findings suggest that dialogue quality most strongly affects fields requiring complex extraction or synthesis from the conversation — particularly allergy history and past medical history — while fields with minimal source information (personal history) remain unaffected, and fields that are uniformly high-modification regardless of input quality (chief complaint, HPI) show limited additional degradation.

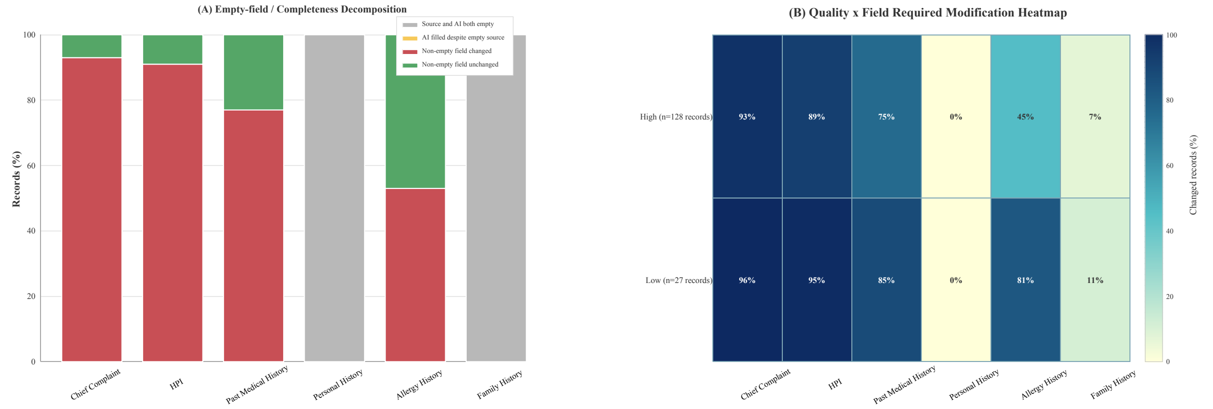

***eFigure 3. Completeness and dialogue-quality analyses for the clinical documentation work flow.*** *(A) Empty-field and completeness decomposition by documentation field. Field status was categorised as source and AI both empty (grey), AI filled despite empty source (orange), non-empty field changed by physician (red), or non-empty field unchanged (green). "Source" refers to extractable information in the source dialogue or physician-reviewed source-derived field. (B) Dialogue-quality-by-field heatmap showing required modification proportions within high-quality (n=123 records) and low-quality (n=27 records) dialogue strata. Counts are shown at the generated-record level because each source dialogue was processed through three pipeline variants.*

#### Appendix.9 Confidence Calibration Detailed Results

Confidence calibration analysis using windowed perplexity over the middle 20%–80% of output tokens showed that GPT-4o's confidence signal was unreliable for clinical triage across languages and settings. For GPT-4o task settings with available token-level traces, AUROC values ranged from 0·41 to 0·66, with the majority near 0·5. ECE ranged from 0·10 to 0·53, far exceeding the ideal threshold of 0·1, indicating systematic overconfidence. The confidence separation gap averaged only 0·003 across all settings, meaning correct and incorrect answers received nearly identical confidence scores.

Low-resource language averages for Ophthalmology 3-type native reasoning were: accuracy 41·3%, ECE 0·443, AUROC 0·543, and top-bottom accuracy gap 0·112. English comparison results were: accuracy 50·7%, ECE 0·310, and AUROC 0·579. For MIMIC-III native reasoning, low-resource average accuracy was 48·7%, ECE 0·219, and AUROC 0·494; English accuracy was 47·8% and AUROC 0·511.

A notable finding emerged for Persian in Ophthalmology 3-type native reasoning: AUROC was highest at 0·658 but ECE was worst at 0·528, with accuracy at only 32·7%. The model's confidence could weakly rank predictions, but assigned approximately 85% confidence to predictions that were only 33% accurate, creating a false sense of reliability.

The AUROC difference between English and low-resource languages was small, 0·02–0·06, indicating that the confidence quality problem is fundamental and not primarily language-specific. Translation pivot reduced ECE consistently, including Ophthalmology 3-type low-resource average ECE from 0·443 under native inference to 0·298 under pivot, paralleling its accuracy improvements. Whether alternative uncertainty estimation approaches, such as semantic entropy or self-consistency, offer better calibration in multilingual medical settings is an important question for future investigation.

The Persian glaucoma setting illustrates why confidence could not be used as a safety filter: AUROC was relatively high, but ECE was poor and accuracy was only 32·7%. The model could weakly rank outputs while still being overconfident.

#### Appendix.10 Subgroup and Fairness Analysis Detailed Results

Using the MIMIC-CXR critical and routine pathology groupings defined in Appendix 3, critical findings showed larger absolute degradation than routine findings. For GPT-4o under native-language reasoning, critical-subset F1 fell from 30·8% in English to 20·0% in Chinese, a 10·8 pp drop, compared with routine-subset F1 falling from 27·9% to 23·0%, a 4·9 pp drop. Back-translation restored critical-subset F1 to 30·4-34·9% across languages and eliminated the critical-routine gap.

High-acuity theme stratification in HealthBench showed asymmetric recovery under translation pivot. For GPT-5.2, overall score gains from back-translation were modest, +4·5 pp in Thai and +5·1 pp in Persian, but emergency-referrals score gains were 2·2–2·4 times larger, +10·1 pp in Thai and +12·2 pp in Persian. The absolute emergency-referrals score under back-translation exceeded the English baseline.

Disease subtype stratification in OphthalmologyEHR-Glaucoma showed that under GPT-4o 3-type, angle-closure glaucoma recall, defined as PACG plus APAC, was 79·3% for English but declined to 68·6% for Chinese and 73·6% for Malay. Under English chain-of-thought, angle-closure recall fell further: Thai reached 66·9% and Chinese 67·8%. Translation pivot restored angle-closure recall to near-baseline levels, 72·7–80·2%. Qwen3 VL showed smaller safety degradation: worst-case angle-closure recall was 81·8% for Thai under English chain-of-thought in the 2-type setting, compared with 85·1% in English.

Prospective documentation workflow subgroup analysis showed that initial visits required 3·8 pp more editing than follow-up visits, 56·3% versus 52·5%. Medium-complexity records had the highest edit burden, 63·9%. Optometry cases required substantially more editing than general eye disease, 63·0% versus 53·0%.

Patient-level demographic stratification by age, gender, and ethnicity was not feasible. Prospective multi-site validation with at least 100 patients per language per disease subtype, stratified by age, gender, and disease severity, was recommended.

#### Appendix.11 Representative Case Studies

##### Appendix.11.1 HealthBench: Language-Conditioned Generation Produces Actively Incorrect Medical Recommendations

The ground-truth expectation for an infant solid-food question is that the model should recommend iron-rich foods as first foods, advocate early and frequent allergen introduction per AAP/LEAP trial evidence, and warn against honey before 12 months due to botulism risk.

In English, the model correctly prioritised iron-rich foods and explicitly advocated early allergen introduction. Thai diverged sharply: the model listed mashed rice or fine rice porridge as the first recommended food, with iron-rich meats appearing only as a secondary mention. This is inconsistent with contemporary infant-feeding guidance that prioritises iron-rich complementary foods and early allergen introduction. The Thai response also framed allergen introduction with excessive caution rather than the evidence-based recommendation of early and frequent exposure. Chinese listed iron-fortified rice cereal among first foods but also prominently recommended meat/liver puree as a high-iron option, partially mitigating the error. Persian triggered three negative items, notably the critical −10 item because the response failed to explain the reason for the honey prohibition, namely risk of infant botulism.

Back-translation partially recovered guideline adherence. Thai-to-English recovered from the rice-cereal error. However, Malay-to-English lost the honey warning that the direct Malay response had retained, illustrating that back-translation can simultaneously introduce its own omissions. The Thai rice-cereal recommendation reflects pre-2016 pediatric feeding guidelines, suggesting that the model’s medical knowledge in Thai draws disproportionately from older training data.

##### Appendix.11.2 MIMIC-CXR: The Same Chest Radiograph Produces Divergent Diagnostic Narratives Across Languages

Subject 12503315, study_id 51367051, is a complex multi-pathology case involving a patient with metastatic adenocarcinoma. CheXpert reference labels confirm six concurrent findings: atelectasis, consolidation, oedema, pleural effusion, pneumothorax, and support devices. Pneumothorax, the indication for the exam, is the highest-priority finding.

English correctly identified the right-sided pneumothorax but missed all other findings. Chinese identified three of six findings, including pneumothorax and atelectasis. The most consequential failures occurred in Malay and Persian. Malay produced an entirely different clinical narrative: it described opacification in the right lung field, diagnosed cardiomegaly and right pleural effusion, but recorded pneumothorax as absent. The model reinterpreted the radiographic signs of pneumothorax as a cardiac-effusion syndrome, a clinically plausible but fundamentally incorrect diagnosis that could lead to substantially different management. Persian explicitly stated that no signs of pneumothorax or rib fracture were observed and constructed a narrative around possible cardiomegaly and left-sided infiltrate.

Back-translation corrected the missed pneumothorax in both Malay and Persian but converged toward a simplified English template that stripped away most other findings. The clinical risk illustrated by this case is language-dependent selective attention to radiographic findings, producing language-conditioned diagnostic divergence. Whether this pattern generalises beyond this individual case requires further systematic evaluation.

##### Appendix.11.3 OphthalmologyEHR-Glaucoma: Multilingual Reasoning Distorts Bilateral Severity Stratification

Patient 0347 had right-eye PACS with cataract and post-LPI, with ground-truth low-risk label, and left-eye PAC with cataract, lens subluxation, and post-SPI, with ground-truth high-risk label. In the 2-type classification, the coding scheme was 1 = PACS, 2 = PAC, 3 = PACG, and 4 = APAC. English classified the right eye as APAC, code 4, representing over-escalation from the ground-truth code 1, and recommended laser peripheral iridotomy. Chinese classified it as PACG, code 3. Malay produced the ground-truth-concordant output.

In the 3-type setting with fundus photographs, the same images were integrated in opposite directions across languages. Persian preserved the right eye as PACS, citing the normal C/D ratio, while English still classified it as APAC. The risk is that severity stratification errors could influence treatment intensity: over-escalation rewrites a milder post-LPI state as APAC requiring urgent intervention, while erroneous downgrading of the left eye recasts progressive PAC as a lower-risk condition.

##### Appendix.11.4 MIMIC-III: Multilingual ICU Reasoning Produces Falsely Reassuring Low-Risk Extubation Narratives

Patient 118 had ground-truth re-intubation within 6 hours labelled Yes. Supporting physiological evidence included $PaO_2/FiO_2$ 87–175, mean 117•5; PEEP persistently 10 $cmH_2O$; pH 7•265–7•31; MAP 52–67; and ongoing dopamine support. Nearly all language–method combinations produced a false-negative prediction of No.

The failure arose at the level of extraction of critical continuous variables. Chinese described pH fluctuations and $PaO_2/FiO_2$ decline as acceptable; Thai stated that the decline was not yet severe enough to suggest respiratory failure. Reassuring cues such as younger a ge and stable RSBI were placed at the centre of the narrative while the features most relevant to extubation safety were downweighted or omitted. Back-translation reduced cross-language variation but did so by converging toward the same milder English intermediate representation, representing shared downshifting of severity rather than recovery of the high-risk physiological state. The potential clinical risk illustrated by this case is selective degradation of actionable physiological parameters, producing a fluent false negative that could, in the absence of clinician review, be mistaken for an acceptable risk assessment. However, this single case should not be interpreted as evidence of a systematic failure pattern without larger-scale validation.

##### Appendix.11.5 Cross-Case Interpretation

Across the representative cases, multilingual degradation may reflect unstable reasoning execution, training-data imbalance, tokenisation differences, prompt alignment, or visual-language integration differences. These mechanisms are plausible but not proven, and the cases should be read as clinical illustrations that support the larger quantitative analyses.

#### Appendix.12 Regulatory and Policy Implications

Current regulatory and governance frameworks for AI in health provide limited explicit guidance on language-specific performance evaluation. The following graded recommendations were proposed.

Supported by direct evidence from both benchmark and prospective evaluation: clinical governance frameworks should retain human oversight for AI-generated medical documentation in all evaluated languages, given the near-universal physician edit rate of 98•7%; regulators, developers, and deploying institutions should consider language-specific safety auditing, including prespecified clinical risk measures beyond aggregate accuracy, for clinical AI systems intended for multilingual use.

Supported by indirect or preliminary evidence, warranting further validation: healthcare institutions may consider implementing language-specific quality-monitoring frameworks, such as the modification-score metric introduced in this study, pending multi-site validation; model developers should report per-language safety profiles alongside aggregate metrics, and the open-source toolkit released with this study provides one practical means of doing so.

The toolkit enables users to inspect exploratory token-level confidence diagnostics, including position-wise perplexity curves, logic-break measures, and confidence-shift visualisations. These diagnostics are intended for auditing and research use and should not be interpreted as validated clinical safety filters.

Taken together, the policy implication is that multilingual deployment should include language-specific safety auditing with prespecified clinical risk endpoints. Modification score may support post-deployment monitoring, but it requires multi-site validation with larger language-specific cohorts.
